# Distinct tissue correlates of proteinuria and hematuria in IgA nephropathy

**DOI:** 10.64898/2026.11.22.26368706

**Authors:** Shaun G. Hong, Ara Ko, Donggun Kim, Hyunah Ku, Jae-Ik Oh, Jinsun Lee, Hajeong Lee, Seung Seok Han, Yaerim Kim, Jinkyung Kwon, Jeonghwan Lee, Jung Pyo Lee, Ji In Park, Jung Tak Park, Jung Nam An, Kyung-Chul Moon, Hyun Je Kim, Young-Gon Kim, Sehoon Park, Dong Ki Kim, the KORNERSTONE investigators

**Author notes:** These authors contributed equally (Shaun G. Hong, Ara Ko). **Corresponding authors** Dong Ki Kim, MD, PhD Professor, Department of Internal Medicine, Seoul National University College of Medicine 101 Daehak-ro, Jongno-gu, Seoul, 03080, Korea, Sehoon Park, MD, PhD Assistant Professor, Department of Internal Medicine, Seoul National University College of Medicine 101 Daehak-ro, Jongno-gu, Seoul, 03080, Korea.

## Abstract

**Background:** Proteinuria, estimated glomerular filtration rate (eGFR) and hematuria guide the care of IgA nephropathy, but hematuria has been less consistently associated with outcome than proteinuria, and whether these measurements reflect the same tissue process is unclear.

**Methods:** Single-cell-level spatial transcriptomics (Xenium, 5,101 targets) profiled 3.08 million cells in biopsies from 60 patients with IgA nephropathy, followed for a median of 8.3 years, and 24 living kidney donors. Injury scores, the proportion of injured glomeruli and glomerular signaling were related to the clinical measurements, to injury in surrounding tubules and to confirmed 40% eGFR decline or kidney failure.

**Results:** Higher proteinuria and lower eGFR were associated with a greater proportion of injured glomeruli, and within biopsies, glomeruli with higher injury scores were surrounded by more injured proximal tubular cells; tubulointerstitial changes were present even with low proteinuria and preserved kidney function. Hematuria was not associated with the injury scores but with higher vascular endothelial growth factor (VEGF) and phosphoinositide 3-kinase and lower p53 and tumor necrosis factor–related apoptosis-inducing ligand signaling in podocytes, changes that were larger with more urinary red blood cells, and glomeruli with higher podocyte VEGF activity were not surrounded by more injured tubular cells. Thirteen patients reached the outcome; the adjusted hazard ratio per 10-percentage-point higher proportion of injured glomeruli was 1.46 (95% confidence interval, 1.12 to 1.90), whereas hematuria and podocyte VEGF activity were not significantly associated.

**Conclusions:** In IgA nephropathy, proteinuria and eGFR accompany established glomerular and tubular injury that is associated with later loss of kidney function, whereas hematuria accompanies podocyte signaling consistent with a pro-survival response.

## Introduction

Proteinuria, estimated glomerular filtration rate (eGFR) and hematuria define the clinical presentation of IgA nephropathy and guide its treatment. The disease develops through mesangial deposition of immune complexes containing galactose-deficient IgA1^1^, and its course ranges from stable kidney function to kidney failure^2^. Proteinuria of 0.5 g/day or more identifies an increased risk of progression, current guidance aims to lower proteinuria below 0.5 and ideally below 0.3 g/day^3^, and remission of proteinuria improves prognosis^4^. The tissue of patients with low proteinuria and preserved eGFR is less well described, and their course has differed between cohorts^5^.

Hematuria has been less consistently associated with outcome. Persistent hematuria^6^ and its remission^7^ have been associated with outcome in some cohorts, the association depends on concurrent proteinuria and on the timing of measurement^8^, and hematuria is not part of the International IgAN Prediction Tool^9^. Red blood cells reach the urine through breaks in the glomerular capillary wall^10,11^, but whether hematuria at biopsy reflects the same glomerular injury as proteinuria or a different glomerular process is unclear.

Tubular atrophy and interstitial fibrosis are among the strongest prognostic features of the Oxford classification^12,13^, and crescents are graded by the proportion of glomeruli involved^14^ because that proportion carries prognostic information^15^. Single-cell and spatial studies have identified inflammatory glomerular changes in mild IgA nephropathy^16^ and injured epithelial states shared across kidney diseases^17,18^. Single-cell-level spatial transcriptomics measures the cellular composition of each glomerulus in a biopsy and of its surrounding tubules, so that glomerular and tubular changes can be compared with the clinical measurements in the same tissue.

We examined whether proteinuria, eGFR and hematuria reflect the same or different tissue processes in IgA nephropathy, which of these processes extends to the surrounding tubules, and which accompanies later loss of kidney function.

## Methods

### Patients and clinical measurements

Archival formalin-fixed, paraffin-embedded kidney biopsies from 60 patients with biopsy-proven IgA nephropathy were obtained from KORNERSTONE, a prospective Korean kidney-biopsy cohort that collects biospecimens with linked clinical data and follow-up^19^. Controls were zero-time biopsies from 24 living kidney donors, obtained at transplantation and processed in the same way. Baseline urine protein-to-creatinine ratio (UPCR) and serum creatinine were the measurements nearest the biopsy date, and eGFR was calculated from creatinine with the 2021 CKD-EPI equation^20^. For description, patients with eGFR of 60 mL/min/1.73 m^2^ or higher were grouped by UPCR below 0.5, 0.5 to below 1, and 1 g/g or more, and patients with eGFR below 60 mL/min/1.73 m^2^ formed a fourth group. Urine red blood cells were taken from the urinalysis recorded at biopsy in the registry, in six microscopy categories (fewer than 1, 1–4, 5–9, 10–49, 50–99 and 100 or more per high-power field); hematuria was defined as five or more red blood cells per high-power field, and the six categories were also analyzed as an ordered scale. Oxford MEST-C scores were taken from the pathology record, which gave no M, E, S or T score for three biopsies taken in 2011. Biopsy tissue was collected with the participants’ written informed consent. The study was approved by the Seoul National University Hospital Institutional Review Board (2511-060-1690).

### Spatial transcriptomics and tissue injury

Single-cell-level spatial transcriptomics (Xenium, 5,101-target panel) profiled 3,083,455 cells from the 84 participants^21^. Cells were assigned to the 24 cell types of the study kidney atlas, including intact and injured proximal tubular cells (Figure 1a). Glomeruli were identified as compact clusters of podocytes, mesangial cells, parietal epithelial cells and juxtaglomerular cells containing at least five podocytes; the patient biopsies contained 627 glomeruli and the donor biopsies 373 (Supplemental Figure 1). In a sensitivity analysis, all clusters were counted regardless of podocyte number.

**Figure 1.**
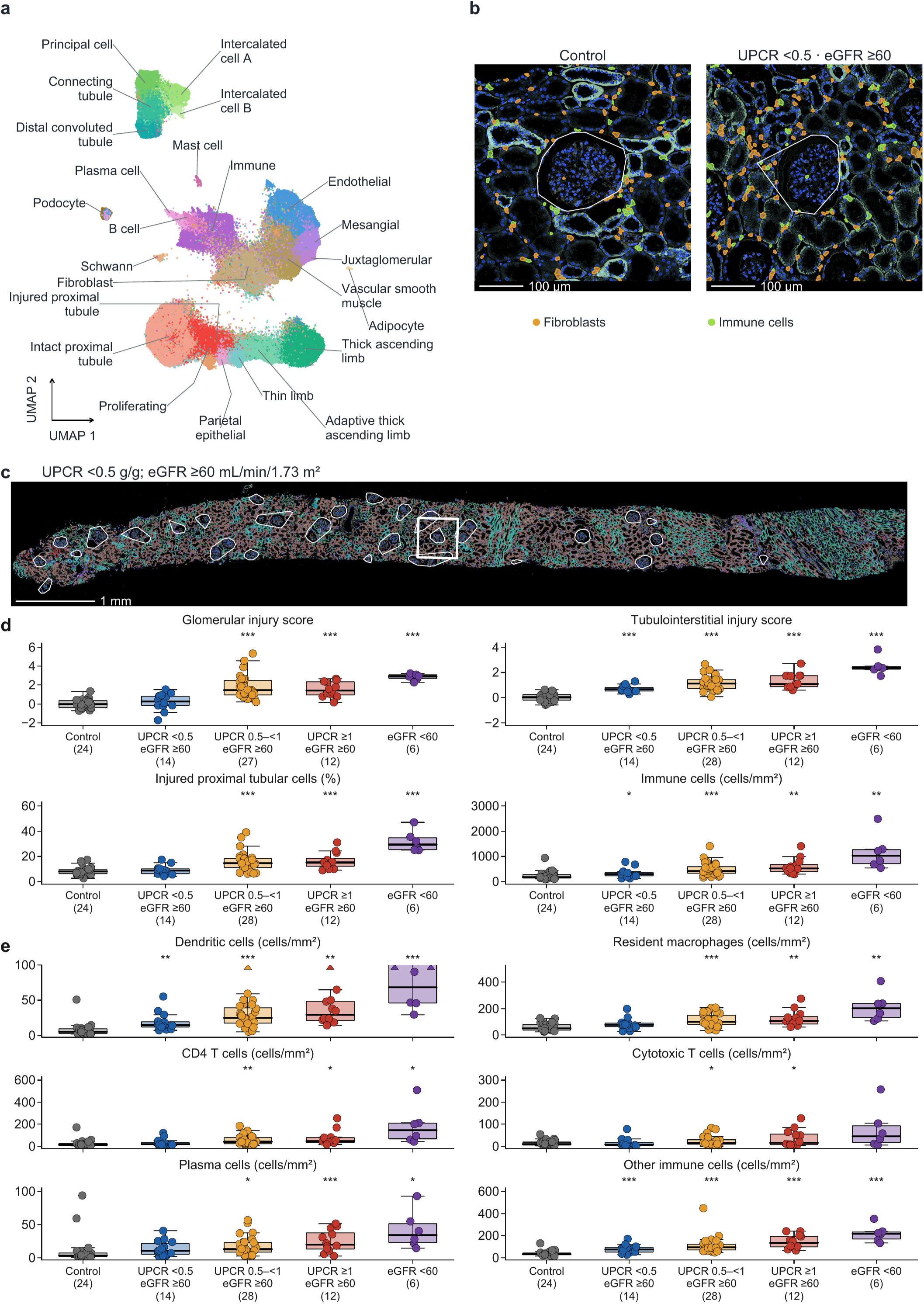
Cell types and tissue injury across clinical groups. (a) Uniform manifold approximation and projection of 385,140 cells sampled from the 24 cell types of the study atlas (up to 20,000 cells per type), colored by cell type and labeled directly; intact and injured proximal tubular cells are shown separately. (b) Fields of 500 by 500 µm from a control kidney and from the low-proteinuria biopsy in (c), each centered on an uninjured glomerulus of typical size and selected as described in the Supplemental Methods, showing the Xenium morphology image with fibroblasts and immune cells filled by type and the central glomerulus outlined in white. Scale bars, 100 µm. (c) Whole section from a patient with proteinuria below 0.5 g/g and eGFR of at least 60 mL/min/1.73 m ^2^, colored as in (a) over the Xenium morphology image; glomeruli are outlined, and the rectangle locates the field shown in (b). Scale bar, 1 mm. (d) Glomerular injury score, tubulointerstitial injury score, proportion of injured proximal tubular cells and immune-cell density in controls (*n* = 24), in patients with eGFR of at least 60 mL/min/1.73 m^2^ and proteinuria below 0.5 g/g (*n* = 14), 0.5 to below 1 g/g (28) or 1 g/g or more (12), and in patients with eGFR below 60 mL/min/1.73 m^2^ (6). Glomerular scores were available in 27 of the 28 patients in the 0.5 to below 1 g/g group. Boxes show medians and interquartile ranges, whiskers extend to observations within 1.5 interquartile ranges, and points are patients; asterisks mark age- and sex-adjusted differences from control (* *P* < 0.05, \*\**P* < 0.01, \*\*\**P* < 0.001). (e) Density of dendritic cells, resident macrophages, CD4 T cells, cytotoxic T cells, plasma cells and other immune cells in the same groups, shown as in (d); triangles mark four dendritic-cell values above 100 cells/mm ^2^. Group sizes beneath each label reflect available measurements. Age- and sex-adjusted differences from control are given in Supplemental Tables 1 and 2. Abbreviations: eGFR, estimated glomerular filtration rate; UPCR, urinary protein-to-creatinine ratio.

For each glomerulus, a glomerular injury score summarized how its cellular composition differed from that of control glomeruli: fewer podocytes, endothelial cells and surrounding tubular cells, and more mesangial, stromal, parietal epithelial, proliferating and immune cells. A tubulointerstitial injury score summarized the tissue around proximal tubules and thick ascending limbs in the same way: fewer intact tubular and endothelial cells, and more fibroblasts and immune cells. Both scores are expressed relative to control kidney, where 0 is the control average and 1 is one control standard deviation; their components and weights are defined in the Supplemental Methods. Higher scores therefore indicate greater loss and gain of cells relative to control kidney. For each patient, the glomerular score was the median across glomeruli and the tubulointerstitial score the mean across tubular sites. Injured proximal tubular cells were those classified as injured in the atlas; in this cohort they expressed more *HAVCR1* and *VCAM1* and less *LRP2* and *WDR72* than intact cells, in keeping with injured tubular states described in human kidney atlases^17,22^. Their proportion among all proximal tubular cells and the density of immune cells per tissue area were calculated for each patient. Immune cells were further classified into dendritic cells, resident macrophages, inflammatory myeloid cells, CD4 and cytotoxic T cells, natural killer-like cells, B cells, plasma cells, neutrophils, mast cells and other immune cells using the immune-cell subtype labels of the study atlas, and the density of each population was calculated.

### Proportion and severity of injured glomeruli

A glomerulus was considered injured when its score exceeded that of 95% of control glomeruli. For each patient, we calculated the proportion of glomeruli that were injured and, among the injured glomeruli, their mean score as the severity of injury. Because severity cannot be calculated in a patient without injured glomeruli, the mean score of the most injured tenth of glomeruli was used as a second measure of severity in all patients with evaluable glomeruli.

### Glomerular signaling programs and surrounding tubules

Gene counts were summed by patient and cell type and normalized^23,24^. The activity of 14 signaling pathways was estimated from the expression of each pathway’s target genes (PROGENy) in podocytes, mesangial cells, glomerular endothelial cells and parietal epithelial cells, and in the immune cells and fibroblasts within 80 µm of proximal tubules^25,26^.

For each glomerulus, the proportion of injured cells among proximal tubular cells in the surrounding tissue (100–300 µm from the glomerulus, outside other glomeruli) was related to its injury score by comparing glomeruli within the same biopsy. The same analysis was repeated with podocyte and mesangial vascular endothelial growth factor (VEGF) activity, scaled to their within-biopsy standard deviations, in place of the injury score. Two further analyses excluded tubular cells adjacent to the glomerulus and restricted the glomerular score to its podocyte, endothelial, mesangial and parietal epithelial components. Spatial definitions and the selection of representative fields are given in the Supplemental Methods.

### Kidney outcome and statistical analysis

The primary outcome was a composite kidney event: an eGFR at or below 60% of the biopsy value (a 40% decline) or below 15 mL/min/1.73 mZ, confirmed by a measurement at least 28 days later or present at the last measurement, or kidney failure, defined as an eGFR below 15 mL/min/1.73 mZ on two measurements; a 40% decline is an accepted surrogate for kidney failure in clinical trials^27,28^. Patients without an event were censored at the last eGFR measurement, and follow-up is reported as the time from biopsy to the last eGFR measurement.

Each tissue measure was related to proteinuria, eGFR and hematuria in one regression model, adjusted for age, sex, acquisition batch and transcript depth; effects are reported per doubling of UPCR, per 10 mL/min/1.73 m^2^ lower eGFR and for hematuria present versus absent. Comparisons with control kidney were adjusted for age and sex. The proportion of injured glomeruli was the primary tissue measure for the kidney-outcome analysis, and the other tissue measures were examined in parallel; each entered a separate Cox model adjusted for baseline eGFR, proteinuria, age and sex. Cumulative incidence across thirds of the proportion of injured glomeruli was compared by the log-rank test and its test for trend. Pathway comparisons were corrected for the 14 pathways within each cell type, and differences with q below 0.10 are reported. Further details are given in the Supplemental Methods.

## Results

### Proteinuria, eGFR and tissue injury

Fourteen patients had low proteinuria and preserved kidney function at biopsy, with proteinuria below 0.5 g/g and eGFR of 60 mL/min/1.73 m^2^ or higher (median 0.34 g/g and 110 mL/min/1.73 m^2^; Table 1). Compared with control kidney, their tubulointerstitial injury score was higher, whereas their glomerular injury score and the proportion of injured proximal tubular cells did not differ significantly (median injured proximal tubular cells, 8.63% versus 7.94% in controls; Figure 1b–d; Supplemental Table 1). Around the tubules, they had fewer intact tubular and endothelial cells and more fibroblasts and immune cells; immune cells around proximal tubules were 1.50 times as dense as in controls, with more dendritic cells and other immune cells (Figure 1e; Supplemental Table 2). Glomerular and tubulointerstitial injury were higher with higher proteinuria and were greatest in patients with eGFR below 60 mL/min/1.73 m^2^ (Figure 1d). With higher proteinuria, Janus kinase–signal transducer and activator of transcription (JAK-STAT) activity was higher in CD4 T cells and dendritic cells around tubules (*q* < 0.10), and tumor necrosis factor α (TNF-α) and nuclear factor κB (NF-κB) activity tended to be higher in fibroblasts and TNF-α activity in inflammatory myeloid cells (unadjusted *P* < 0.05; Supplemental Figure 2; Supplemental Data 1).

**Table 1.** Clinical characteristics.

| Characteristic | Control kidney | UPCR <0.5, eGFR ≥60 | UPCR 0.5–<1, eGFR ≥60 | UPCR ≥1, eGFR ≥60 | eGFR <60 |
|---|---|---|---|---|---|
| Patients, *n* | 24 | 14 | 28 | 12 | 6 |
| Female / male, *n* | 17 / 7 | 8 / 6 | 12 / 16 | 6 / 6 | 4 / 2 |
| Age, years | 55 (50–58) | 38 (29–43) | 41 (33–48) | 36 (33–43) | 56 (48–58) |
| eGFR, mL/min/1.73 m² | 103 (99–106) | 110 (91–119) | 92 (82–108) | 106 (89–117) | 52 (51–54) |
| UPCR, g/g | 0.05 (0.03–0.06) | 0.34 (0.23–0.37) | 0.82 (0.64–0.93) | 3.42 (3.19–3.98) | 2.69 (1.19–4.04) |
| Urine red blood cells <1/HPF, *n* | 13 | 1 | 0 | 1 | 0 |
| Urine red blood cells 1–4/HPF, *n* | 11 | 1 | 4 | 0 | 0 |
| Urine red blood cells 5–9/HPF, *n* | 0 | 1 | 2 | 0 | 0 |
| Urine red blood cells 10–49/HPF, *n* | 0 | 7 | 16 | 7 | 4 |
| Urine red blood cells 50–99/HPF, *n* | 0 | 2 | 2 | 1 | 1 |
| Urine red blood cells ≥100/HPF, *n* | 0 | 2 | 4 | 3 | 1 |
| Hematuria (≥5/HPF), *n* | 0 | 12 | 24 | 11 | 6 |
| Oxford M1, *n*/*N* (%) | — | 1/12 (8) | 5/27 (19) | 7/12 (58) | 1/6 (17) |
| Oxford E1, *n*/*N* (%) | — | 3/12 (25) | 3/27 (11) | 2/12 (17) | 3/6 (50) |
| Oxford S1, *n*/*N* (%) | — | 3/12 (25) | 21/27 (78) | 9/12 (75) | 5/6 (83) |
| Oxford T1–2, *n*/*N* (%) | — | 0/12 (0) | 1/27 (4) | 3/12 (25) | 2/6 (33) |
| Oxford C1–2, *n*/*N* (%) | — | 2/14 (14) | 3/28 (11) | 3/12 (25) | 1/6 (17) |
| Evaluable glomeruli per biopsy | 13 (9–21) | 11 (8–16) | 9 (5–12) | 10 (6–14) | 12 (10–15) |
| Follow-up, years | — | 8.29 (6.53–10.17) | 8.18 (7.14–9.44) | 9.08 (8.08–10.84) | 7.63 (6.72–8.63) |
| 40% eGFR decline or kidney failure, *n*/*N* | — | 0/14 | 8/28 | 3/12 | 2/6 |

Values are median (interquartile range), n or n/N (%). UPCR is g/g; eGFR is mL/min/1.73 m², calculated with the 2021 CKD-EPI equation. Controls are living kidney donors, and their values are from the evaluation before donation. Follow-up is the time from biopsy to the last eGFR measurement. Kidney failure denotes eGFR below 15 mL/min/1.73 m² on two measurements. Oxford percentages are of the patients with a score; the pathology record gave no M, E, S or T score for three biopsies taken in 2011. All 11 patients with E1 and all nine with C1–2 had hematuria. Abbreviations: C, crescents; CKD-EPI, Chronic Kidney Disease Epidemiology Collaboration; E, endocapillary hypercellularity; eGFR, estimated glomerular filtration rate; HPF, high-power field; M, mesangial hypercellularity; S, segmental glomerulosclerosis; T, tubular atrophy and interstitial fibrosis; UPCR, urinary protein-to-creatinine ratio.

In models that included proteinuria, eGFR and hematuria together, higher proteinuria was associated with both the glomerular and the tubulointerstitial injury score, and lower eGFR with the glomerular score (Table 2).

**Table 2.** Tissue measures in relation to proteinuria, eGFR and hematuria.

| Tissue measure | Per doubling of UPCR | Per 10 mL/min/1.73 m<sup>2</sup> lower eGFR | Hematuria present versus absent |
|---|---|---|---|
| Glomerular injury score | 0.37 (0.18 to 0.57) | 0.25 (0.11 to 0.39) | −0.09 (−0.88 to 0.69) |
| Tubulointerstitial injury score | 0.15 (0.04 to 0.26) | 0.17 (0.02 to 0.31) | 0.13 (−0.34 to 0.60) |
| Injured proximal tubular cells (%) | 1.83 (0.24 to 3.41) | 1.91 (−0.02 to 3.84) | 0.24 (−9.23 to 9.70) |
| Immune-cell density (cells/mm<sup>2</sup>) | 66.32 (−18.29 to 150.93) | 70.98 (−40.30 to 182.26) | 61.63 (−319.02 to 442.28) |
| Proportion of injured glomeruli (%) | 9.78 (5.67 to 13.89) | 5.12 (1.73 to 8.51) | −7.96 (−40.46 to 24.55) |
| Severity of injury in injured glomeruli | 0.158 (−0.001 to 0.316) | 0.164 (0.010 to 0.318) | −0.065 (−1.238 to 1.108) |

Estimates (95% CI) from one joint model per tissue measure, adjusted for age, sex, acquisition batch and transcript depth. Hematuria is ≥5 red blood cells/HPF. All 60 patients were included, except 59 for the glomerular injury score and the proportion of injured glomeruli, because one biopsy had no evaluable glomeruli, and 52 for the severity of injury, which was assessed in patients with at least one injured glomerulus. Abbreviations: CI, confidence interval; eGFR, estimated glomerular filtration rate; HPF, high-power field; UPCR, urinary protein-to-creatinine ratio.

The median proportion of injured glomeruli was 4.71% in control kidney, 11.90% in patients with low proteinuria and preserved kidney function, 42.86% and 56.25% in those with preserved kidney function and proteinuria of 0.5 to below 1 g/g and of at least 1 g/g, and 82.91% in those with eGFR below 60 mL/min/1.73 m^2^ (Figure 2a,b; Supplemental Table 3).

**Figure 2.**
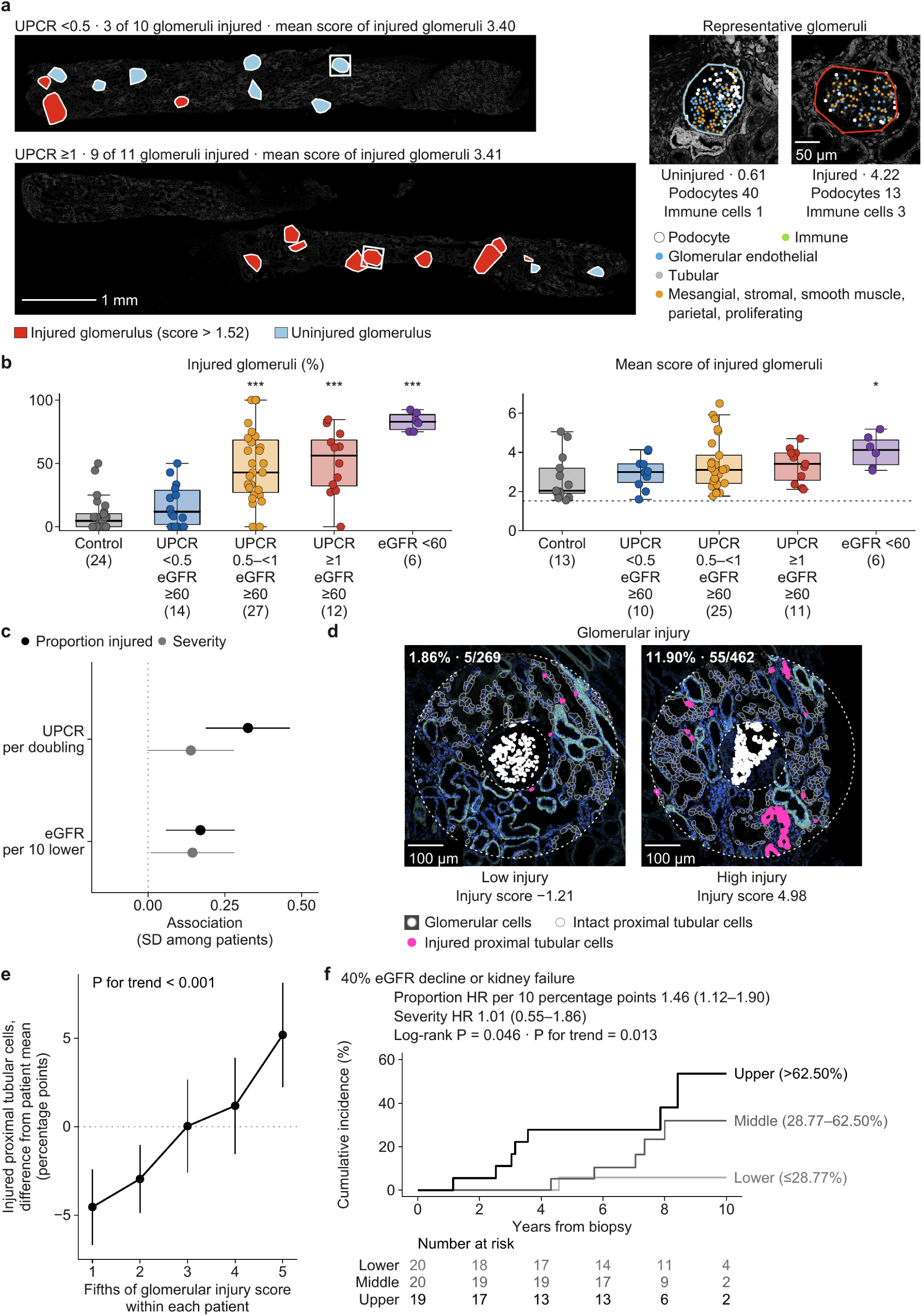
Proportion of injured glomeruli, surrounding tubules and later decline in kidney function. (a) Whole sections from two patients with mean scores of 3.40 and 3.41 among their injured glomeruli and a lower and a higher proportion of injured glomeruli (3 of 10 and 9 of 11). Each glomerulus is drawn over the Xenium morphology image as its boundary enlarged 1.5-fold for visibility, in red if injured and in blue if not. Insets at right show representative uninjured and injured glomeruli, with the numbers of podocytes and immune cells within each glomerular boundary; white squares locate them in the sections. Podocytes, glomerular endothelial cells, mesangial, stromal and parietal epithelial cells, and immune cells are colored by group, and tubular cells are shown in gray, over the grayscale morphology image. The whole sections share the same spatial scale and a 1-mm scale bar; inset scale bar, 50 µm. (b) Proportion of injured glomeruli (left) and mean score of the injured glomeruli (right) in controls and in the clinical groups of Figure 1d; group sizes are given beneath each label, and the right panel includes participants with at least one injured glomerulus. Boxes, whiskers and points are as in Figure 1d, and asterisks mark age- and sex-adjusted differences from control (* *P* < 0.05, \*\**P* < 0.01, \*\*\**P* < 0.001). (c) Associations of proteinuria per doubling and of eGFR per 10 mL/min/1.73 m^2^ lower with the proportion of injured glomeruli (black) and with the severity of their injury (gray), from the models of Table 2, expressed in standard deviations of each measure among patients, with 95% confidence intervals. (d) Two glomeruli from one biopsy with a low and a high glomerular injury score. Fields show the Xenium morphology image with glomerular cells in white; within the ring 100–300 µm from the glomerular center, injured proximal tubular cells are filled in magenta and intact proximal tubular cells outlined in gray, and tissue outside the ring is dimmed. The number and percentage of injured cells among proximal tubular cells in the ring are given in each field, and the injury score beneath each field. Scale bar, 100 µm. (e) Injured proximal tubular cells in the surrounding ring, grouped into within-patient fifths of glomerular injury score, in 49 patients with at least five evaluable glomeruli; values are differences from each patient’s mean, points and vertical lines show means and 95% confidence intervals across patients, and the *P* value is for linear trend across fifths. (f) Cumulative incidence of confirmed 40% eGFR decline or kidney failure by thirds of the proportion of injured glomeruli ( *n* = 59; cut points 28.77% and 62.50%), with numbers at risk, the log-rank test, the test for trend and the adjusted hazard ratios for the proportion and the severity of injured glomeruli (Table 3). Abbreviations: CI, confidence interval; eGFR, estimated glomerular filtration rate; HR, hazard ratio; UPCR, urinary protein-to-creatinine ratio.

**Table 3.** Tissue measures and 40% decline in eGFR or kidney failure.

| Tissue measure | Adjusted hazard ratio (95% CI) | *P* |
|---|---|---|
| Proportion of injured glomeruli (per 10 percentage points) | 1.46 (1.12 to 1.90) | 0.005 |
| Severity of injury in injured glomeruli | 1.01 (0.55 to 1.86) | 0.971 |
| Mean score of the most injured tenth of glomeruli | 1.10 (0.86 to 1.40) | 0.466 |
| Median glomerular injury score | 1.77 (1.09 to 2.89) | 0.022 |
| Tubulointerstitial injury score | 1.86 (0.59 to 5.89) | 0.293 |
| Injured proximal tubular cells (per 10 percentage points) | 1.45 (0.67 to 3.11) | 0.343 |
| Immune-cell density (per 100 cells/mm²) | 0.95 (0.76 to 1.19) | 0.677 |
| Podocyte VEGF activity | 0.82 (0.48 to 1.40) | 0.470 |
| Mesangial VEGF activity | 1.30 (0.65 to 2.62) | 0.458 |
| Hematuria (present versus absent) | 0.52 (0.11 to 2.57) | 0.425 |

Each tissue measure entered a separate Cox model adjusted for baseline eGFR, ln(UPCR), age and sex. The proportion of injured glomeruli was the primary tissue measure; the remaining measures were examined in parallel. Thirteen of 60 patients reached the outcome, and all 13 were included in every model. All 60 patients were included, except 59 for the proportion of injured glomeruli, the mean score of the most injured tenth, the median glomerular injury score and podocyte VEGF activity, and 52 for the severity of injury in injured glomeruli. Proportions use ten percentage points, immune density 100 cells/mm², and scores and pathway activities one unit. Abbreviations: CI, confidence interval; eGFR, estimated glomerular filtration rate; UPCR, urinary protein-to-creatinine ratio; VEGF, vascular endothelial growth factor.

Across patients, each doubling of proteinuria was associated with 9.78 percentage points more injured glomeruli (95% confidence interval [CI], 5.67 to 13.89), and each 10 mL/min/1.73 m^2^ lower eGFR with 5.12 percentage points more (1.73 to 8.51). The severity of injury within injured glomeruli was also higher with both, by a similar amount: 0.158 score units per doubling of proteinuria (−0.001 to 0.316; *P* = 0.051) and 0.164 per 10 mL/min/1.73 m^2^ lower eGFR (0.010 to 0.318; *P* = 0.038); expressed in standard deviations among patients, the association of proteinuria with the proportion of injured glomeruli was more than twice that with severity (Figure 2c; Table 2). Counting all clusters gave similar associations (Supplemental Table 3). Higher proteinuria and lower eGFR therefore accompanied more widespread glomerular injury.

### Injured glomeruli and surrounding tubules

Within the same biopsy, glomeruli with a higher injury score were surrounded by more injured proximal tubular cells. For each one-unit higher injury score, the proportion of injured cells among the surrounding proximal tubular cells was 2.00 percentage points higher (95% CI, 0.97 to 3.04; 623 glomeruli in 56 patients); between the least and most injured fifths of glomeruli within a biopsy, the proportion differed by 9.73 percentage points, in the same direction in every clinical group (Figure 2d,e; Supplemental Table 4). The result was similar when tubular cells adjacent to the glomerulus were excluded (1.92 percentage points; 0.85 to 2.98) and when the score was restricted to its glomerular components (1.86; 0.71 to 3.00) (Supplemental Table 4).

### Hematuria and podocyte signaling

Fifty-three patients had hematuria and seven did not. After accounting for proteinuria and eGFR, the glomerular and tubulointerstitial injury scores, the proportion of injured proximal tubular cells, immune-cell density and the proportion and severity of injured glomeruli did not differ with hematuria (Table 2; Figure 3a), and unadjusted comparisons gave the same results (Supplemental Table 5).

**Figure 3.**
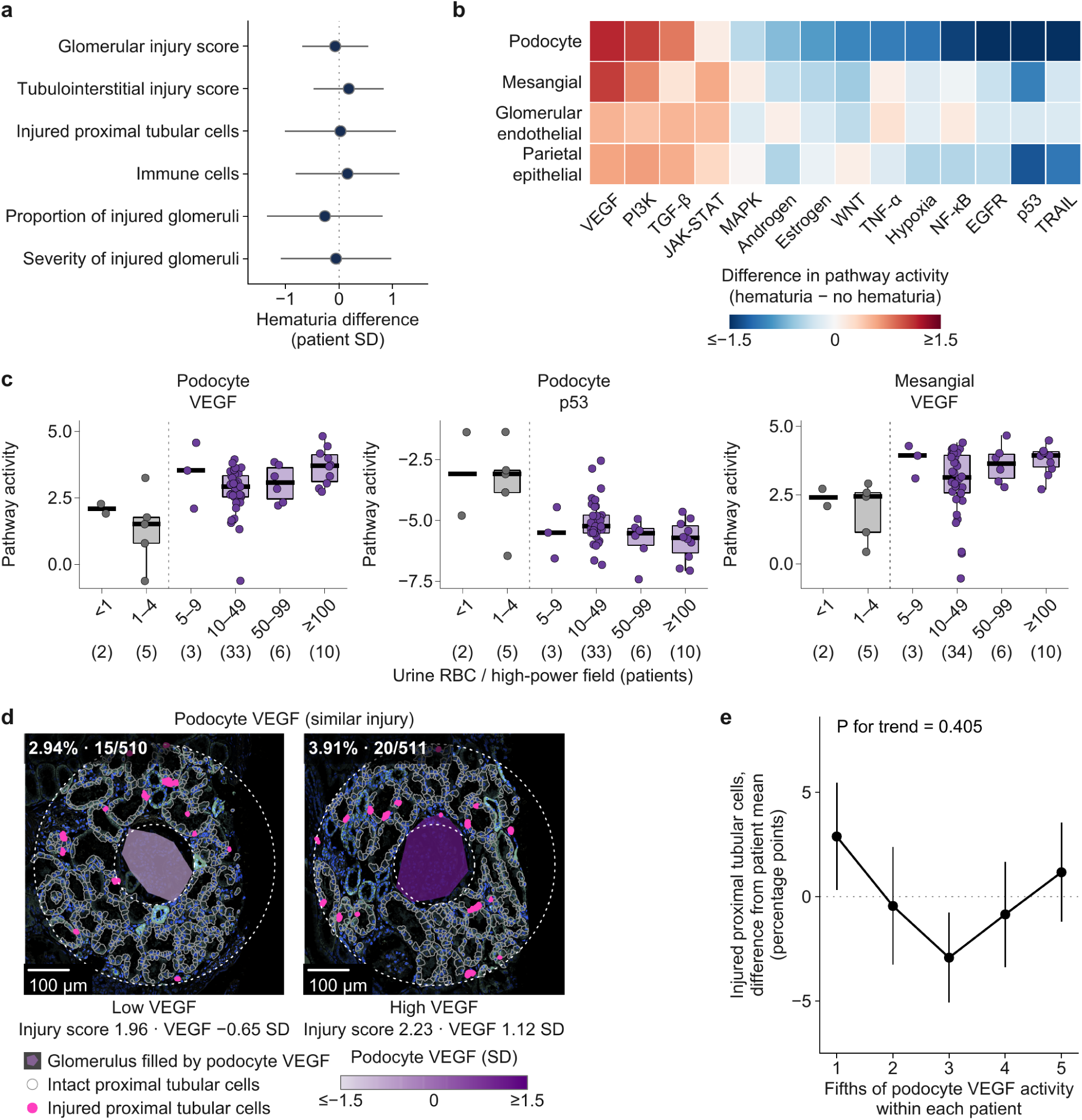
Hematuria, podocyte signaling and surrounding tubules. (a) Differences between patients with and without hematuria in six tissue injury measures, including the injury scores, adjusted for proteinuria, eGFR, age, sex, acquisition batch and transcript depth, each expressed in standard deviations of the measure among patients, with 95% confidence intervals. (b) Differences in the activity of 14 signaling pathways in podocytes, mesangial cells, glomerular endothelial cells and parietal epithelial cells between patients with and without hematuria, adjusted as in (a); pathways are ordered by the difference in podocytes, and *q* values are given in Supplemental Table 6. (c) Podocyte VEGF, podocyte p53 and mesangial VEGF pathway activity across the six urine microscopy categories (*n* = 2, 5, 3, 33, 6 and 10 for podocyte measures and 2, 5, 3, 34, 6 and 10 for mesangial VEGF); categories below the hematuria threshold of 5 red blood cells per high-power field are gray, boxes show medians and interquartile ranges for categories with more than three patients, and points are patients; categories with three or fewer patients show points and the median. (d) Two glomeruli from one biopsy with similar injury scores and a low and a high podocyte VEGF activity, displayed as in Figure 2d; each glomerulus is filled according to its podocyte VEGF activity, and podocyte VEGF activity in within-biopsy standard deviations is given beneath each field. Scale bar, 100 µm. (e) Injured proximal tubular cells in the surrounding ring, grouped into within-patient fifths of podocyte VEGF activity, displayed as in Figure 2e. Abbreviations: eGFR, estimated glomerular filtration rate; EGFR, epidermal growth factor receptor; HPF, high-power field; JAK-STAT, Janus kinase–signal transducer and activator of transcription; MAPK, mitogen-activated protein kinase; NF-κB, nuclear factor κB; PI3K, phosphoinositide 3-kinase; TGF-β, transforming growth factor β; TNF-α, tumor necrosis factor α; TRAIL, tumor necrosis factor–related apoptosis-inducing ligand; UPCR, urinary protein-to-creatinine ratio; VEGF, vascular endothelial growth factor.

Hematuria was instead associated with signaling programs of podocytes and mesangial cells. Podocytes of patients with hematuria had higher VEGF and phosphoinositide 3-kinase (PI3K) activity and lower p53, tumor necrosis factor–related apoptosis-inducing ligand (TRAIL), hypoxia, epidermal growth factor receptor (EGFR), NF-κB and estrogen activity, and mesangial cells had higher VEGF activity; glomerular endothelial and parietal epithelial cells showed smaller changes, mostly in the same directions (Figure 3b; Supplemental Table 6). Across the six microscopy categories, podocyte VEGF and PI3K and mesangial VEGF activity increased, and podocyte p53 and TRAIL activity decreased, with the number of urinary red blood cells, whereas the injury scores did not change across categories (Figure 3c; Supplemental Table 5). Hematuria therefore marked a glomerular signaling program rather than the higher injury scores that accompanied proteinuria and reduced eGFR.

Within biopsies, the proportion of injured surrounding proximal tubular cells did not differ with podocyte VEGF activity (−0.58 percentage points per within-biopsy standard deviation; 95% CI, −2.26 to 1.10; 623 glomeruli in 56 patients) or with mesangial VEGF activity (−1.17; −3.07 to 0.72; 606 glomeruli in 56 patients), and showed no linear trend across fifths of podocyte VEGF activity within each biopsy (Figure 3d,e; Supplemental Table 4).

### Proportion of injured glomeruli, hematuria and decline in kidney function

During a median follow-up of 8.3 years, 13 of 60 patients reached a confirmed 40% decline in eGFR or kidney failure. Cumulative incidence rose across thirds of the proportion of injured glomeruli, with 1, 5 and 7 events in the lower, middle and upper thirds (log-rank *P* = 0.046; *P* for trend = 0.013; Figure 2f). After adjustment for baseline eGFR, proteinuria, age and sex, the hazard ratio per 10-percentage-point higher proportion of injured glomeruli was 1.46 (95% CI, 1.12 to 1.90; *P* = 0.005), whereas that for the severity of injury within injured glomeruli was 1.01 (0.55 to 1.86). The median glomerular injury score of each biopsy, which is higher when more of its glomeruli are injured, was also associated with the outcome (1.77; 1.09 to 2.89). Hematuria at biopsy, glomerular VEGF activity, the tubulointerstitial injury score (1.86; 0.59 to 5.89), injured proximal tubular cells (1.45; 0.67 to 3.11) and immune-cell density were not significantly associated with the outcome (Table 3). Counting all clusters gave a similar result (Supplemental Table 3).

## Discussion

In IgA nephropathy, proteinuria and eGFR reflected one process in the kidney and hematuria another. Higher proteinuria and lower eGFR were associated with a greater proportion of injured glomeruli, injured glomeruli were surrounded by injured tubules, and the proportion of injured glomeruli was associated with later loss of kidney function. Hematuria was associated with podocyte signaling consistent with a pro-survival response, without the established glomerular and tubular injury linked to loss of kidney function.

For injury defined by cellular composition, the present biopsies showed the pattern known for crescents in the Oxford classification, in which the fraction of glomeruli involved is associated with outcome^14,15^: higher proteinuria and lower eGFR were associated with a greater proportion of injured glomeruli, whereas the severity of injury within injured glomeruli varied less with proteinuria and was not associated with later decline. Podocyte depletion links glomerular injury to glomerulosclerosis^29^. Proteinuria is also a cause of tubular injury: filtered protein activates inflammatory pathways in proximal tubular cells^30,31^, and tubular responses to glomerular proteinuria have been described in kidney tissue^32^. The peritubular capillaries downstream of an injured glomerulus also supply the surrounding interstitium^33^. Because injured glomeruli were surrounded by injured tubules, a larger number of injured glomeruli could involve a larger volume of surrounding tubulointerstitium, which may explain the association between the proportion of injured glomeruli and later loss of kidney function.

Hematuria implies damage to the glomerular capillary wall^10,11^. The injury scores used here measure changes in cellular composition that accompany established injury, such as loss of podocytes, endothelial cells and surrounding tubular cells, and in these biopsies they were associated with reduced eGFR and later loss of kidney function. They did not differ with hematuria once proteinuria and eGFR were considered. Hematuria was instead associated with higher VEGF and PI3K and lower p53, TRAIL and NF-κB signaling in podocytes. Podocyte-derived VEGF maintains the glomerular endothelium^34^, and both its loss^35,36^ and its excess^37^ produce glomerular disease, whereas p53 and TRAIL promote podocyte death^38,39^. These changes suggest a pro-survival podocyte response in patients with hematuria, without the cell loss that accompanies established injury. Every patient with endocapillary hypercellularity or crescents had hematuria (Table 1), consistent with damage to the capillary wall in these active lesions.

Tubular atrophy and interstitial fibrosis are among the strongest histological predictors of kidney prognosis^12,13^, and the hematuria-associated signaling was not accompanied by injury in the surrounding tubules. This separation between capillary-wall damage with a podocyte response and established glomerular and tubulointerstitial injury may explain why the association of hematuria with outcome has varied between cohorts^6–8^ and why hematuria is not part of the International IgAN Prediction Tool^9^. Remission of hematuria during treatment may therefore reflect subsiding glomerular activity rather than recovery from established injury.

Changes around tubules were present even in patients with low proteinuria and preserved kidney function, with more fibroblasts and immune cells and a proportion of injured tubular cells similar to that in controls. Immune and fibroblast responses occur with injured epithelial states in kidney atlases^17,18^ and spatial studies^40,41^, after acute injury^42,43^ and in fibrosis^44^. Current guidance identifies risk of progression from proteinuria of 0.5 g/day^3^, patients with minimal proteinuria have generally had favorable long-term courses^5^, and the treatments that lower proteinuria and preserve kidney function, corticosteroids^45^, targeted-release budesonide^46^ and sparsentan^47^, were tested in patients with substantially higher proteinuria. Trials in patients whose proteinuria is already low would show whether these peritubular changes respond to treatment.

This study has several limitations. Donor controls were older than the patients and were obtained at transplantation; comparisons with controls were adjusted for age and sex. Hematuria was assessed from a single registry urinalysis recorded at biopsy, and few patients had no hematuria. The proportion and severity of injured glomeruli were measured in one tissue section, and the surrounding tubules were defined in two dimensions rather than as connected nephrons. Pathway activities were inferred from target-gene expression on a targeted panel. Treatment during follow-up was not included in the outcome models, and the number of outcome events was modest.

In IgA nephropathy, proteinuria and eGFR accompany established injury in glomeruli and their surrounding tubules that is associated with later loss of kidney function, whereas hematuria accompanies podocyte signaling consistent with a pro-survival response.

## Acknowledgments

We thank all participants, donors and KORNERSTONE investigators. The biospecimens and data used for this study were provided by the Biobank of Seoul National University Hospital, a member of the Korea Biobank Network, and by the Korea National Institute of Health (KNIH) research project (project No. KBN5_A03).

## Author Contributions

S.G.H. performed the spatial and statistical analyses and drafted the manuscript. A.K. curated the clinical data, interpreted the results and drafted the manuscript. D.K. performed cell-type annotation of the spatial transcriptomic data. H.K., J.-I.O., J.L., H.L., S.S.H., Y.K., J.K., J.H.L., J.P.L., J.I.P., J.T.P., J.N.A., K.-C.M., H.J.K. and Y.-G.K. contributed to participant enrollment and data generation and reviewed the final manuscript. S.P. conceived the study, curated the clinical data, drafted the manuscript and supervised the analyses. D.K.K. established the cohort and supervised the study. All authors revised the manuscript and approved the final version.

## Data Sharing Statement

Individual-level spatial transcriptomic and clinical data are not publicly available because the archival biopsy tissue was collected under consent for research in Korea, and neither the consent nor the approval of the Institutional Review Board of Seoul National University Hospital covers transfer of individual-level data outside Korea. De-identified data can be shared through a collaboration agreement with the KORNERSTONE investigators, subject to approval by the Institutional Review Board and the Biobank of Seoul National University Hospital; requests should be addressed to the corresponding authors. Summary statistics are provided in Supplemental Tables 1–6 and Supplemental Data 1.

