## Supplemental information for "Distinct tissue correlates of proteinuria and hematuria in IgA nephropathy"

#### Table of Contents

**Supplemental Methods** 2
Clinical measurements 2
Glomerular and tubulointerstitial injury scores 2
Proportion and severity of injured glomeruli 4
Glomerular signaling programs 4
Associations with proteinuria, eGFR and hematuria 5
Glomerular injury and the surrounding tubules 5
Kidney outcome 6
Representative fields 6
Spatial image display 7

**Supplemental Results** 8
Injured proximal tubular cells and low-proteinuria tissue changes 8
Patients with at least five glomeruli 8

**Supplemental Tables** 9
Supplemental Table 1. Tissue measures by clinical group 9
Supplemental Table 2. Immune-cell density by location and population 10
Supplemental Table 3. Proportion and severity of injured glomeruli 11
Supplemental Table 4. Glomeruli and their surrounding tubules 12
Supplemental Table 5. Amount of hematuria 13
Supplemental Table 6. Glomerular signaling programs and hematuria 14

**Supplemental Figures** 15
Supplemental Figure 1. Identification of glomeruli and the injury threshold 15
Supplemental Figure 2. Proteinuria and signaling in cells around proximal tubules 16

**Supplemental Data 1. Proteinuria and signaling around proximal tubules (separate Excel file)**

#### Supplemental Methods

##### Clinical measurements

Serum creatinine values (mg/dL) measured on the same date were summarized by their median. The value nearest the biopsy date was taken as baseline, and the earlier value was used when two dates were equally close; all baseline values fell within nine days of the biopsy. Baseline UPCR was selected by the same rule. eGFR was calculated from creatinine with the 2021 CKD-EPI equation without race, using sex and the age in decimal years on each measurement date, counted from the first day of the birth month, for the baseline and for every subsequent measurement[1]. Measurements before adulthood, on or after kidney-replacement therapy, or outside 1–250 mL/min/1.73 $m^2$ were excluded. No clinical value was imputed.

##### Glomerular and tubulointerstitial injury scores

Glomeruli were identified from the coordinates of podocytes, mesangial cells, parietal epithelial cells and juxtaglomerular cells. Density-based clustering used a 60-µm neighborhood and at least five anchor cells to form a core; clusters with at least ten anchor cells were eligible, and those containing at least five podocytes were retained for the primary analysis. The convex hull of the anchor cells defined the glomerular boundary, and their mean coordinate defined its center. Clustering and boundaries were calculated separately within each tissue section. The patient biopsies contained 627 glomeruli and the donor biopsies 373. Analyses were performed in Python with scikit-learn, SciPy and NumPy.

For each anchor cell, we counted all other cells within 20, 40 and 80 µm and calculated the fraction of each of the 24 atlas cell types among those neighbors, excluding the anchor itself. For glomerulus $g$, $p_{gart}$ denotes this fraction averaged over the anchor cells of class $a$, at radius $r$, for neighboring cell type $t$. Each represented anchor class contributed equally, irrespective of its abundance. When an anchor class was absent from a glomerulus, the average was taken over the anchor classes present. Let $A_g$ denote the anchor classes present in glomerulus $g$.

For a quantity $x$, donor standardization was

$$Z_C(x) = \frac{x - \mu_C(x)}{\sigma_C(x)}.$$

Here $\mu_C$ and $\sigma_C$ are the mean and sample standard deviation of the specified donor reference. For each neighborhood feature, the reference comprised retained donor glomeruli in which the corresponding anchor class was present.

Six components described reduced podocyte representation; reduced endothelial representation; increased mesangial, stromal and vascular smooth-muscle representation; increased parietal epithelial and proliferating-cell representation; increased immune-cell representation, including B, plasma and mast cells; and reduced tubular representation. The tubular component included intact and injured proximal tubule, conventional and adaptive thick ascending limb, descending thin limb, distal convoluted and connecting tubules, and collecting-duct principal and intercalated cells. For component $k$, let $T_k$ be its member cell types and let $s_k$ be −1 for the three reduced-cell components and +1 for the three increased-cell components. Components and the final score were calculated as

$$C_{gk} = Z_C\left[s_k \,\mathrm{mean}_{a\in A_g,\, r\in\{20,40,80\},\, t\in T_k}\left\{Z_C(p_{gart})\right\}\right],$$

$$G_g = Z_C\left(\frac{1}{6}\sum_{k=1}^{6} C_{gk}\right).$$

Thus, cell types within a component, the anchor classes present, and radii received equal weights at the first averaging step; each of the six standardized components then received weight 1/6. All three standardization steps used the pooled 373 retained donor glomeruli. A participant's glomerular score was the median across his or her glomeruli. A glomerulus was injured when its score exceeded the donor 95th percentile, 1.52. The four-component sensitivity score retained the podocyte, endothelial, mesangial/stromal and parietal epithelial/proliferating components and applied the same final standardization.

The all-cluster sensitivity analysis included all 572 donor and 901 patient clusters, irrespective of podocyte number. For that analysis, absent-anchor fractions were replaced by their corresponding pooled donor means, all four anchor classes were averaged, and donor standardization and the 95th-percentile threshold of 1.75 used all 572 donor clusters.

The tubulointerstitial score used the neighborhoods of proximal tubular cells, including injured cells, and thick ascending limb cells, including adaptive cells. Up to 1,500 cells of each lineage per section were sampled without replacement. At 40 and 80 µm, four grouped fractions described intact tubular cells, stromal and vascular smooth-muscle cells, immune cells including B, plasma and mast cells, and endothelial cells. Intact tubular cells included all tubular classes listed above except injured proximal tubule and adaptive thick ascending limb. The index cell was excluded from both numerator and denominator. For tubular site $j$ of lineage $\ell$, with grouped fraction $p_{jrk}$, the signed component and final score were

$$u_{jk} = \frac{s_k}{2}\sum_{r\in\{40,80\}} p_{jrk},$$

$$T_j = Z_C\left[\frac{1}{4}\sum_{k=1}^{4} Z_{C,\ell}(u_{jk})\right].$$

The sign was negative for intact tubular and endothelial cells and positive for stromal and immune cells. Component standardization used donor sites of the same proximal-tubular or thick-ascending-limb lineage; the equally weighted four-component mean was then standardized against all sampled donor sites. A participant's tubulointerstitial score was the mean across his or her sampled sites. The donor glomeruli and tubular sites therefore each had a score mean of zero and sample standard deviation of one; the participant summaries were not standardized again.

Injured and intact proximal tubular cells were the cells classified as injured proximal tubule and as proximal tubule in the 24-class study atlas. The injured-cell proportion was the number of injured cells divided by the number of intact plus injured proximal tubular cells. To characterize these states, observed mean transcripts per cell for *HAVCR1*, *VCAM1*, *LRP2* and *WDR72* were compared within each patient, requiring at least 20 cells of each state.

Occupied tissue area was the number of 50-by-50-µm tiles containing at least one cell, multiplied by 0.0025 $mm^2$. Immune-cell density was the number of cells of all immune classes (including B, plasma and mast cells) divided by this area. Glomerular regions for immune-density measurements included all clusters formed from the pooled cells of the four glomerular anchor classes, without the podocyte-count restriction. The requirement of at least five podocytes applied to the glomerular injury score and its associated local tubular analyses. Location-specific densities were measured within the glomerular regions, within 50 µm outside their boundaries, within 50 µm of proximal tubular cells outside those regions, within 50 µm of endothelial or vascular smooth-muscle cells outside those regions, and in the remaining interstitium. The remaining interstitium excluded the other four locations. Each location used its occupied tissue area as denominator; locations may overlap.

#### Proportion and severity of injured glomeruli

The injury threshold was the 95th percentile of the 373 retained donor glomerular scores (1.52). For each patient, the proportion of injured glomeruli was the number of glomeruli above this threshold divided by the number of glomeruli identified, and the severity of injury was the mean score of the glomeruli above the threshold. One patient had no qualifying glomerulus and seven other patients had no glomerulus above the threshold, leaving 59 proportions and 52 severities. The mean score of the most injured tenth of glomeruli (all glomeruli at or above the patient's 90th percentile, ties retained) was defined for all 59 patients. A sensitivity analysis required at least five identified glomeruli.

#### Glomerular signaling programs

Transcript counts were summed by participant and cell type, requiring at least 20 cells, and normalized by the trimmed mean of M values with log2 counts per million (prior count 2) across all 5,101 targets[2,3]. Pathway activity was estimated with the 14-pathway PROGENy model as the univariate linear fit of normalized expression on the pathway's gene weights across all panel genes, requiring at least five panel-measured target genes; the activity score is the t statistic of that fit[4,5]. Participant-level pseudobulks required at least 20 cells and were formed for immune populations in whole tissue and within 80 µm of proximal tubular cells, and for fibroblasts within 80 µm. For the glomerulus-level comparison, counts from podocytes or mesangial cells within each glomerular boundary were summed. Glomerulus-level counts were normalized together with those of all glomeruli containing that cell type. VEGF activity was estimated on that scale, and the spatial analyses used the resulting values for glomeruli meeting the podocyte criterion.

#### Associations with proteinuria, eGFR and hematuria

Each tissue measure or pathway activity was regressed on log2 UPCR, eGFR divided by −10, hematuria (five or more red blood cells per high-power field), age, sex, acquisition batch and the natural logarithm of one plus mean transcripts per cell, so that the coefficients correspond to a doubling of UPCR, 10 mL/min/1.73 $m^2$ lower eGFR and hematuria present versus absent. Sensitivity analyses replaced the hematuria indicator with thresholds of 10, 50 or 100 red blood cells per high-power field, or with the six microscopy categories scored 0 to 5; the ordinal coefficient describes one category step. Comparisons of each clinical group with controls, and of adjacent groups, used separate two-group regressions adjusted for age and sex, because all donor biopsies belonged to one acquisition batch. Available cases were used without imputation. Linear models used heteroskedasticity-robust (HC3) standard errors with t-based confidence intervals[6]. Pathway comparisons were corrected for the 14 pathways within each cell type by the Benjamini–Hochberg procedure[7], and differences with $q$ below 0.10 are reported. Immune-population densities were compared between the low-proteinuria group and controls with adjustment for age and sex, and related to the three clinical measurements in the same joint clinical model.


#### Glomerular injury and the surrounding tubules

For each glomerulus, the surrounding tubules were the proximal tubular cells 100–300 µm from the glomerular centroid and outside every glomerular boundary meeting the podocyte criterion, and the outcome was the percentage of injured cells among them; 626 glomeruli in 59 patients had at least one surrounding proximal tubular cell. Three patients had only one evaluable glomerulus and contributed no within-patient comparison, leaving 623 glomeruli in 56 informative patients. The single-predictor model related this percentage to the glomerular injury score with a separate intercept for each patient, so that only differences among a patient's own glomeruli contributed. The same model was fitted with podocyte or mesangial VEGF activity, centered and scaled within patient, as the predictor. Each patient's glomeruli were weighted to contribute equally. Standard errors were clustered by patient (CR1 correction) with t-based intervals[8]. One sensitivity analysis excluded tubular cells within 80 µm of any anchor cell of the index glomerulus, and another used the four-component glomerular score with the same exclusion. For the display by fifths, the 49 patients with at least five evaluable glomeruli and complete podocyte VEGF values contributed patient-centered outcomes, averaged within fifths of each glomerular measure with equal weight per patient. Trends across within-patient fifths in Figures 2e and 3e were tested with the fifth as a continuous term in a linear model with patient-specific intercepts.

#### Kidney outcome

Follow-up eGFR was recalculated from every measured creatinine with the same equation as the baseline. The primary outcome was a composite kidney event: the first date on which eGFR was at or below 60% of the baseline (a 40% decline) or below 15 mL/min/1.73 m² and this was confirmed by a measurement at least 28 days later or was the last measurement, or kidney failure (eGFR below 15 mL/min/1.73 m² on two measurements), whichever came first. Patients without an event were censored at their last eGFR measurement, and follow-up is reported as the time from biopsy to that measurement.

Each tissue measure entered a separate Cox model with baseline eGFR, ln UPCR, age and sex, with Efron's handling of ties[9]; the proportion of injured glomeruli was the primary tissue measure and the others were examined in parallel. Hazard ratios are per 10 percentage points for proportions, per 100 cells/mm$^2$ for immune density and per unit for scores and pathway activities. Proportional hazards were checked with scaled Schoenfeld residuals[10]; for the proportion of injured glomeruli in the primary model, the predictor-specific test gave $P$=0.601 and the global test $P$=0.198. Cumulative incidence was shown as one minus the Kaplan–Meier estimate[11] across thirds of the proportion of injured glomeruli (cut points 28.77% and 62.50%; 20, 20 and 19 patients), compared by the three-group log-rank test and by its test for trend with scores 1, 2 and 3[12].

#### Representative fields

The control and low-proteinuria fields in Figure 1b were selected from 500 by 500 µm fields centered on uninjured glomeruli whose boundary area lay within the interquartile range of all 1,000 evaluable glomeruli, with at least 90% of the field inside the tissue footprint. Fibroblast and immune-cell densities were calculated per tissue area in each field. Each density was centered on its group median among fields meeting the tissue-coverage criterion and divided by the corresponding group interquartile range; the field with the smallest Euclidean distance from the two group medians was selected. For Figure 2a, the two biopsies were selected from patients with at least three injured glomeruli and a mean injured score within 0.5 units of their median, as the biopsies nearest the median proportion in the lower and in the upper third of these patients. Representative uninjured and injured glomeruli for the Figure 2a insets were chosen among glomeruli whose boundary area lay within the interquartile range of all evaluable patient glomeruli and whose complete boundary lay within the tissue, as the uninjured glomerulus of the lower-proportion biopsy and the injured glomerulus of the higher-proportion biopsy whose six component values were closest to the mean component values of all uninjured and all injured patient glomeruli, respectively. Both 275 by 275 µm fields were centered on the bounding rectangles of their actual glomerular boundaries. In the insets, glomeruli were drawn with their actual boundaries. In the whole sections, the convex hulls were enlarged 1.5-fold about their centers for display only and colored according to whether their scores exceeded the injury threshold. The two whole sections were displayed at the same physical scale. For Figure 2d, all within-patient pairs from the lowest and highest injury-score fifths were considered among patients with at least five evaluable glomeruli. Both 100–300-µm rings had to lie at least 80% within the tissue footprint, and both glomerular boundary areas had to lie within the interquartile range of all 1,000 evaluable glomeruli. The pair whose difference in injured proximal tubular cells was closest to the mean within-biopsy difference between the least and most injured fifths (9.73 percentage points) was selected. The tissue footprint was the area covered by occupied 50-by-50-µm tiles, with internal gaps included. For Figure 3d, eligible biopsies had at least five complete fields and a pair differing by no more than 0.5 injury-score units and by at least one within-biopsy standard deviation of VEGF activity; both rings had to lie at least 80% within the tissue footprint, and both glomerular boundary areas had to lie within the same interquartile range. Among these biopsies, the biopsy nearest the median patient-specific injury-score–tubular-injury slope was selected. Within that biopsy, the eligible VEGF pair nearest the median paired difference in surrounding tubular injury was shown.

#### Spatial image display

Xenium morphology images were registered to cell boundaries and transcript coordinates using the recorded pixel size of 0.2125 µm. DAPI was displayed in blue, the ATP1A1/CD45/E-cadherin channel in pale gray-green, and the available 18S and alpha-smooth-muscle-actin/vimentin channels at lower intensity. Channel intensities were scaled between the 1st and 99.5th percentiles of the registered overview for each selected section; the same limits were used for all fields from that section. Selected cell types were shown using their measured segmentation polygons, except in the Figure 2a insets, where filled points marked measured cell centroids within or on the glomerular boundary at full opacity, with identical colors and point sizes in both fields. In Figure 1b, fibroblasts and immune cells were displayed with 60% fill and colored outlines. In Figures 2d and 3d, injured proximal tubular cells were filled in magenta and intact proximal tubular cells were outlined in gray; tissue outside the 100–300-µm ring was dimmed with a 60% black overlay. In Figure 3d, each glomerular boundary was filled according to the podocyte VEGF activity of that glomerulus, centered and scaled within the biopsy, on a light-to-dark purple scale spanning −1.5 to +1.5 standard deviations.

### Supplemental Results

#### Injured proximal tubular cells and low-proteinuria tissue changes

Across paired comparisons in 60 patients, injured proximal tubular cells had more transcripts per cell for *HAVCR1* and *VCAM1* and fewer for *LRP2* and *WDR72* than intact cells. The age- and sex-adjusted differences between patients with proteinuria below 0.5 g/g and controls are given in Supplemental Table 1.

Among immune populations, other immune cells accounted for the largest part of the increase in the low-proteinuria group. With adjustment for age and sex, dendritic-cell density was higher by 13.54 cells/mm$^2$ (95% CI, 4.93 to 22.16) in the low-proteinuria group than in controls. Resident macrophages were higher by 29.69 cells/mm$^2$ (95% CI, −0.24 to 59.63) and other immune cells by 67.73 cells/mm$^2$ (30.49 to 104.97; Supplemental Table 2).

#### Patients with at least five glomeruli

In the 49 patients with at least five glomeruli, each doubling of UPCR was associated with 9.79 percentage points more injured glomeruli (95% CI, 5.45 to 14.14) and each 10 mL/min/1.73 m$^2$ lower eGFR with 5.28 (2.01 to 8.55). For severity ($n$ = 45), the corresponding estimates were 0.10 score units (−0.07 to 0.26) and 0.09 (−0.04 to 0.22).

### Supplemental Tables

#### Supplemental Table 1. Tissue measures by clinical group

| Measure | Clinical group | Median (interquartile range) | Adjusted difference from control (95% CI) |
|---|---|---|---|
| Glomerular injury score | Control kidney | −0.03 (−0.39–0.34) | Reference |
| Glomerular injury score | UPCR <0.5, eGFR ≥60 | 0.25 (−0.16–0.83) | 0.46 (−0.22 to 1.14) |
| Glomerular injury score | UPCR 0.5–<1, eGFR ≥60 | 1.46 (0.97–2.49) | 1.79 (1.08 to 2.50) |
| Glomerular injury score | UPCR ≥1, eGFR ≥60 | 1.41 (0.96–2.33) | 1.93 (1.33 to 2.52) |
| Glomerular injury score | eGFR <60 | 2.91 (2.71–3.06) | 2.86 (2.46 to 3.26) |
| Tubulointerstitial injury score | Control kidney | 0.03 (−0.20–0.24) | Reference |
| Tubulointerstitial injury score | UPCR <0.5, eGFR ≥60 | 0.68 (0.52–0.80) | 0.88 (0.59 to 1.18) |
| Tubulointerstitial injury score | UPCR 0.5–<1, eGFR ≥60 | 1.11 (0.74–1.42) | 1.35 (1.07 to 1.63) |
| Tubulointerstitial injury score | UPCR ≥1, eGFR ≥60 | 1.09 (0.87–1.74) | 1.85 (1.23 to 2.47) |
| Tubulointerstitial injury score | eGFR <60 | 2.38 (2.27–2.47) | 2.49 (1.79 to 3.18) |
| Injured proximal tubular cells (%) | Control kidney | 7.94 (6.18–9.53) | Reference |
| Injured proximal tubular cells (%) | UPCR <0.5, eGFR ≥60 | 8.63 (6.28–10.14) | 2.26 (−0.17 to 4.69) |
| Injured proximal tubular cells (%) | UPCR 0.5–<1, eGFR ≥60 | 14.63 (11.11–18.41) | 8.07 (3.83 to 12.32) |
| Injured proximal tubular cells (%) | UPCR ≥1, eGFR ≥60 | 15.14 (12.21–18.57) | 13.10 (6.35 to 19.84) |
| Injured proximal tubular cells (%) | eGFR <60 | 29.34 (25.49–34.62) | 23.06 (14.20 to 31.92) |
| Immune-cell density (cells/mm²) | Control kidney | 179.55 (138.54–286.34) | Reference |
| Immune-cell density (cells/mm²) | UPCR <0.5, eGFR ≥60 | 299.20 (229.82–360.38) | 198.08 (29.50 to 366.65) |
| Immune-cell density (cells/mm²) | UPCR 0.5–<1, eGFR ≥60 | 417.46 (336.34–586.94) | 319.28 (169.41 to 469.14) |
| Immune-cell density (cells/mm²) | UPCR ≥1, eGFR ≥60 | 524.26 (399.30–677.60) | 650.64 (252.90 to 1048.38) |
| Immune-cell density (cells/mm²) | eGFR <60 | 1020.51 (719.84–1268.93) | 915.06 (306.85 to 1523.27) |
| Reduced preserved tubular representation | UPCR <0.5 versus control | — | 0.50 (0.29 to 0.72) |
| Stromal representation | UPCR <0.5 versus control | — | 0.84 (0.56 to 1.12) |
| Immune representation | UPCR <0.5 versus control | — | 0.43 (0.10 to 0.75) |
| Reduced endothelial representation | UPCR <0.5 versus control | — | 0.30 (0.09 to 0.52) |

Comparisons are adjusted for age and sex. Component rows describe the four contributions to tubulointerstitial injury in patients with UPCR <0.5 g/g versus controls.

#### Supplemental Table 2. Immune-cell density by location and population

| Location | Control median | UPCR <0.5 median | Adjusted difference (95% CI) | *P* |
|---|---|---|---|---|
| Within glomeruli | 91.02 | 150.30 | 112.17 (−10.39 to 234.74) | 0.072 |
| Glomerular border | 212.41 | 287.18 | 127.41 (−7.46 to 262.29) | 0.063 |
| Around proximal tubules | 175.96 | 264.59 | 145.44 (15.68 to 275.19) | 0.029 |
| Around vessels | 176.38 | 300.98 | 199.19 (29.76 to 368.63) | 0.023 |
| Remaining interstitium | 61.00 | 96.73 | 113.35 (−115.94 to 342.63) | 0.322 |

**Immune-cell populations**

| Population | Control median | UPCR <0.5 median | Adjusted difference (95% CI) | *P* | Per doubling of UPCR (95% CI) |
|---|---|---|---|---|---|
| Dendritic cells | 4.98 | 14.42 | 13.54 (4.93 to 22.16) | 0.003 | 6.25 (0.96 to 11.55) |
| Resident macrophages | 50.50 | 77.21 | 29.69 (−0.24 to 59.63) | 0.052 | 9.85 (−1.82 to 21.52) |
| Inflammatory myeloid cells | 39.74 | 44.26 | 9.71 (−9.40 to 28.83) | 0.309 | 7.29 (−2.61 to 17.19) |
| CD4 T cells | 13.63 | 19.81 | 25.15 (−8.10 to 58.40) | 0.134 | 7.96 (−10.97 to 26.89) |
| Cytotoxic T cells | 10.59 | 7.96 | 4.50 (−8.56 to 17.56) | 0.489 | 3.04 (−6.09 to 12.17) |
| Natural killer-like cells | 4.83 | 4.74 | 5.18 (−3.53 to 13.89) | 0.235 | 1.44 (−2.44 to 5.32) |
| B cells | 2.56 | 5.24 | 13.53 (−2.86 to 29.92) | 0.103 | 0.69 (−9.10 to 10.48) |
| Plasma cells | 3.23 | 10.49 | 10.77 (−0.24 to 21.78) | 0.055 | 2.90 (−0.95 to 6.74) |
| Neutrophils | 5.13 | 3.95 | 0.89 (−2.45 to 4.22) | 0.592 | 1.02 (−0.06 to 2.11) |
| Mast cells | 1.00 | 2.55 | 2.19 (0.38 to 4.01) | 0.019 | 1.25 (0.32 to 2.18) |
| Other immune cells | 31.11 | 73.00 | 67.73 (30.49 to 104.97) | <0.001 | 21.34 (8.33 to 34.35) |

Density is cells/mm$^2$; spatial regions in the first block may overlap. Population densities in the second block use the whole captured tissue area. Immune populations follow the immune-cell subtype labels of the study atlas; cells without an assigned population are 'other immune cells'. Low-proteinuria comparisons are adjusted for age and sex; associations among patients use the joint clinical model of Table 2.

#### Supplemental Table 3. Proportion and severity of injured glomeruli

| Clinical group | Patients, *n* | Glomeruli per biopsy | Injured glomeruli (%) | Severity in injured glomeruli | Most injured tenth mean score |
|---|---|---|---|---|---|
| Control kidney | 24 | 13 (9–21) | 4.71 (0.00–10.44) | 2.04 (1.90–3.20) | 1.42 (0.93–2.22) |
| UPCR <0.5, eGFR ≥60 | 14 | 11 (8–16) | 11.90 (1.79–28.75) | 2.99 (2.47–3.41) | 2.42 (1.29–4.43) |
| UPCR 0.5–<1, eGFR ≥60 | 28 | 9 (5–12) | 42.86 (27.08–68.33) | 3.11 (2.42–3.85) | 4.00 (2.79–6.28) |
| UPCR ≥1, eGFR ≥60 | 12 | 10 (6–14) | 56.25 (32.14–68.33) | 3.41 (2.57–3.97) | 4.69 (3.59–6.26) |
| eGFR <60 | 6 | 12 (10–15) | 82.91 (76.70–88.50) | 4.13 (3.36–4.64) | 7.31 (5.24–9.10) |

**Clinical associations**

| Measure | Per doubling of UPCR | Per 10 lower eGFR | Hematuria present versus absent |
|---|---|---|---|
| Mean score of the most injured tenth of glomeruli | 0.60 (0.16 to 1.04) | 0.40 (−0.10 to 0.91) | 0.70 (−1.58 to 2.99) |
| All clusters, regardless of podocyte content | | | |
| Proportion of injured glomeruli (%) | 7.97 (5.34 to 10.59) | 3.80 (1.45 to 6.15) | 1.98 (−12.56 to 16.53) |
| Severity of injury in injured glomeruli | 0.34 (0.07 to 0.61) | 0.06 (−0.14 to 0.26) | −0.34 (−1.41 to 0.73) |

**All clusters, regardless of podocyte content: kidney outcome**

| Tissue measure | Adjusted hazard ratio (95% CI) | *P* | Events / patients |
|---|---|---|---|
| Proportion of injured glomeruli (per 10 percentage points) | 1.69 (1.15 to 2.50) | 0.008 | 13/59 |

Values are median (interquartile range). The main analysis includes clusters with at least five podocytes: 373 donor and 627 patient glomeruli; injury exceeds the donor 95th percentile (1.52). Severity is unavailable when no glomerulus exceeds this threshold. The all-cluster sensitivity retains 572 donor and 901 patient clusters, with standardization and the 95th percentile (1.75) recalculated in all 572 donor clusters. Outcome models adjust for baseline eGFR, ln(UPCR), age and sex. One patient in the UPCR 0.5–<1 group had no qualifying glomerulus.

#### Supplemental Table 4. Glomeruli and their surrounding tubules

| Predictor | Population or field definition | Difference in injured tubular cells (95% CI) | Glomeruli / patients | *P* |
|---|---|---|---|---|
| Glomerular injury score (per score unit) | All patients | 2.00 (0.97 to 3.04) | 623 / 56 | <0.001 |
| | UPCR <0.5, eGFR ≥60 | 0.89 (−0.83 to 2.62) | 177 / 13 | 0.282 |
| | UPCR 0.5–<1, eGFR ≥60 | 2.77 (1.11 to 4.42) | 252 / 26 | 0.002 |
| | UPCR ≥1, eGFR ≥60 | 2.39 (0.30 to 4.48) | 116 / 11 | 0.029 |
| | eGFR <60 | 0.58 (−2.25 to 3.41) | 78 / 6 | 0.619 |
| | Excluding adjacent tubular cells | 1.92 (0.85 to 2.98) | 623 / 56 | <0.001 |
| | Glomerular components only | 1.86 (0.71 to 3.00) | 623 / 56 | 0.002 |
| Podocyte VEGF (per within-patient SD) | All glomeruli | −0.58 (−2.26 to 1.10) | 623 / 56 | 0.490 |
| | Excluding adjacent tubular cells | −0.54 (−2.22 to 1.13) | 623 / 56 | 0.516 |
| Mesangial VEGF (per within-patient SD) | All glomeruli | −1.17 (−3.07 to 0.72) | 606 / 56 | 0.220 |
| | Excluding adjacent tubular cells | −1.25 (−3.17 to 0.66) | 606 / 56 | 0.196 |

Differences are percentage points of injured cells among the surrounding proximal tubular cells, per one unit of glomerular injury score or per one within-patient standard deviation of VEGF activity. Within-patient models use equal patient weights and patient-clustered intervals.

#### Supplemental Table 5. Amount of hematuria

| Measure | Hematuria present versus absent, unadjusted | Per microscopy category | ≥10 versus <10/HPF | ≥50 versus <50/HPF | ≥100 versus <100/HPF |
|---|---|---|---|---|---|
| Glomerular injury score | 0.17 (−0.95 to 1.29) *P* = 0.763 | 0.01 (−0.20 to 0.21) *P* = 0.939 | 0.02 (−0.62 to 0.66) *P* = 0.943 | 0.04 (−0.53 to 0.61) *P* = 0.890 | −0.08 (−0.78 to 0.62) *P* = 0.828 |
| Tubulointerstitial injury score | 0.19 (−0.29 to 0.67) *P* = 0.440 | 0.04 (−0.07 to 0.16) *P* = 0.433 | 0.11 (−0.27 to 0.49) *P* = 0.568 | 0.09 (−0.26 to 0.44) *P* = 0.613 | 0.00 (−0.45 to 0.46) *P* = 0.990 |
| Injured proximal tubular cells (%) | 0.46 (−9.19 to 10.12) *P* = 0.924 | 0.19 (−1.90 to 2.28) *P* = 0.853 | 1.35 (−5.51 to 8.20) *P* = 0.695 | 0.60 (−4.60 to 5.81) *P* = 0.817 | −1.68 (−8.60 to 5.24) *P* = 0.628 |
| Immune-cell density (cells/mm²) | 102.46 (−270.63 to 475.55) *P* = 0.585 | 30.55 (−42.32 to 103.42) *P* = 0.404 | 72.24 (−205.63 to 350.11) *P* = 0.604 | 63.75 (−155.67 to 283.17) *P* = 0.562 | 14.30 (−271.37 to 299.96) *P* = 0.920 |
| Proportion of injured glomeruli (%) | −1.69 (−36.42 to 33.05) *P* = 0.923 | −1.55 (−8.69 to 5.59) *P* = 0.665 | −3.87 (−27.45 to 19.71) *P* = 0.743 | −1.01 (−15.45 to 13.43) *P* = 0.889 | −3.89 (−22.27 to 14.50) *P* = 0.673 |
| Severity of injury in injured glomeruli | 0.14 (−1.13 to 1.42) *P* = 0.821 | 0.13 (−0.15 to 0.41) *P* = 0.346 | 0.18 (−0.57 to 0.94) *P* = 0.623 | 0.40 (−0.29 to 1.09) *P* = 0.248 | 0.34 (−0.57 to 1.25) *P* = 0.452 |
| Podocyte VEGF | 1.43 (0.40 to 2.46) *P* = 0.007 | 0.36 (0.13 to 0.58) *P* = 0.002 | 0.67 (−0.26 to 1.59) *P* = 0.153 | 0.71 (0.20 to 1.23) *P* = 0.008 | 0.76 (0.30 to 1.21) *P* = 0.002 |
| Podocyte PI3K | 1.33 (0.15 to 2.51) *P* = 0.028 | 0.35 (0.12 to 0.59) *P* = 0.004 | 0.56 (−0.32 to 1.44) *P* = 0.208 | 0.78 (0.28 to 1.28) *P* = 0.003 | 0.66 (0.17 to 1.15) *P* = 0.010 |
| Podocyte p53 | −1.90 (−3.42 to −0.38) *P* = 0.015 | −0.56 (−0.91 to −0.21) *P* = 0.002 | −1.33 (−2.54 to −0.12) *P* = 0.032 | −0.89 (−1.62 to −0.16) *P* = 0.018 | −0.61 (−1.40 to 0.18) *P* = 0.127 |
| Podocyte TRAIL | −2.47 (−4.41 to −0.52) *P* = 0.014 | −0.62 (−1.06 to −0.19) *P* = 0.006 | −1.62 (−3.08 to −0.16) *P* = 0.030 | −0.95 (−1.91 to 0.01) *P* = 0.052 | −0.43 (−1.49 to 0.62) *P* = 0.415 |
| Podocyte hypoxia | −1.06 (−1.80 to −0.32) *P* = 0.006 | −0.22 (−0.42 to −0.01) *P* = 0.039 | −0.69 (−1.36 to −0.02) *P* = 0.042 | −0.09 (−0.52 to 0.33) *P* = 0.664 | 0.05 (−0.50 to 0.59) *P* = 0.866 |
| Mesangial VEGF | 1.19 (0.38 to 2.00) *P* = 0.005 | 0.31 (0.11 to 0.51) *P* = 0.003 | 0.57 (−0.15 to 1.30) *P* = 0.120 | 0.66 (0.14 to 1.19) *P* = 0.014 | 0.45 (−0.18 to 1.08) *P* = 0.161 |

Estimates (95% CI). The first comparison is unadjusted; all remaining comparisons are adjusted for proteinuria, eGFR, age, sex, acquisition batch and transcript depth in the joint clinical models. The six microscopy categories are scored 0–5. Confidence intervals and *P* values use heteroskedasticity-consistent (HC3) standard errors and the residual t distribution. Hematuria was defined as ≥5 red blood cells per high-power field.

#### Supplemental Table 6. Glomerular signaling programs and hematuria

| Pathway | Podocytes ($n$ = 59) | Mesangial cells ($n$ = 60) | Glomerular endothelial cells ($n$ = 60) | Parietal epithelial cells ($n$ = 60) |
|---|---|---|---|---|
| Androgen | −0.66 (−1.70 to 0.39) $q$ = 0.236 | −0.37 (−0.89 to 0.16) $q$ = 0.384 | 0.09 (−0.20 to 0.39) $q$ = 0.621 | −0.46 (−1.09 to 0.17) $q$ = 0.431 |
| EGFR | −1.58 (−2.83 to −0.34) $q$ = 0.048 | −0.34 (−1.15 to 0.47) $q$ = 0.783 | −0.25 (−0.97 to 0.46) $q$ = 0.611 | −0.40 (−1.37 to 0.57) $q$ = 0.636 |
| Estrogen | −0.85 (−1.72 to 0.02) $q$ = 0.095 | −0.44 (−0.93 to 0.05) $q$ = 0.219 | −0.24 (−0.53 to 0.06) $q$ = 0.316 | −0.11 (−1.33 to 1.12) $q$ = 0.927 |
| Hypoxia | −1.09 (−1.77 to −0.41) $q$ = 0.011 | −0.21 (−0.85 to 0.43) $q$ = 0.783 | −0.16 (−0.54 to 0.21) $q$ = 0.595 | −0.45 (−1.19 to 0.29) $q$ = 0.453 |
| JAK-STAT | 0.12 (−1.33 to 1.56) $q$ = 0.872 | 0.57 (−0.94 to 2.09) $q$ = 0.783 | 0.49 (−0.39 to 1.36) $q$ = 0.564 | 0.31 (−0.67 to 1.30) $q$ = 0.736 |
| MAPK | −0.41 (−1.08 to 0.25) $q$ = 0.236 | 0.12 (−0.47 to 0.70) $q$ = 0.800 | −0.21 (−0.59 to 0.17) $q$ = 0.564 | 0.03 (−0.78 to 0.84) $q$ = 0.946 |
| NF-κB | −1.40 (−2.79 to −0.01) $q$ = 0.095 | −0.07 (−1.09 to 0.95) $q$ = 0.889 | 0.14 (−0.61 to 0.90) $q$ = 0.705 | −0.45 (−1.52 to 0.63) $q$ = 0.636 |
| PI3K | 1.05 (0.07 to 2.03) $q$ = 0.084 | 0.71 (0.04 to 1.39) $q$ = 0.219 | 0.44 (−0.11 to 0.98) $q$ = 0.316 | 0.63 (−0.27 to 1.53) $q$ = 0.431 |
| TGF-β | 0.79 (−0.39 to 1.98) $q$ = 0.236 | 0.20 (−0.69 to 1.10) $q$ = 0.800 | 0.51 (−0.12 to 1.14) $q$ = 0.316 | 0.56 (−0.06 to 1.17) $q$ = 0.353 |
| TNF-α | −1.05 (−2.28 to 0.19) $q$ = 0.132 | 0.09 (−0.79 to 0.97) $q$ = 0.889 | 0.23 (−0.39 to 0.85) $q$ = 0.611 | −0.21 (−1.25 to 0.83) $q$ = 0.877 |
| TRAIL | −2.46 (−3.97 to −0.96) $q$ = 0.011 | −0.28 (−1.23 to 0.67) $q$ = 0.783 | −0.22 (−1.14 to 0.70) $q$ = 0.679 | −1.08 (−2.10 to −0.07) $q$ = 0.259 |
| VEGF | 1.16 (0.14 to 2.18) $q$ = 0.075 | 1.07 (0.37 to 1.77) $q$ = 0.049 | 0.52 (0.01 to 1.03) $q$ = 0.316 | 0.60 (−0.29 to 1.49) $q$ = 0.431 |
| WNT | −0.98 (−2.04 to 0.07) $q$ = 0.104 | −0.54 (−1.12 to 0.04) $q$ = 0.219 | −0.39 (−0.88 to 0.09) $q$ = 0.316 | 0.10 (−0.69 to 0.89) $q$ = 0.927 |
| p53 | −2.01 (−3.28 to −0.75) $q$ = 0.011 | −1.03 (−2.17 to 0.12) $q$ = 0.219 | −0.25 (−0.78 to 0.29) $q$ = 0.595 | −1.29 (−2.27 to −0.31) $q$ = 0.152 |

Hematuria-positive minus negative differences (95% CI) adjust for UPCR, eGFR, age, sex, acquisition batch and transcript depth. $q$ is Benjamini–Hochberg FDR across 14 pathways per cell population.

### Supplemental Figures

#### Supplemental Figure 1

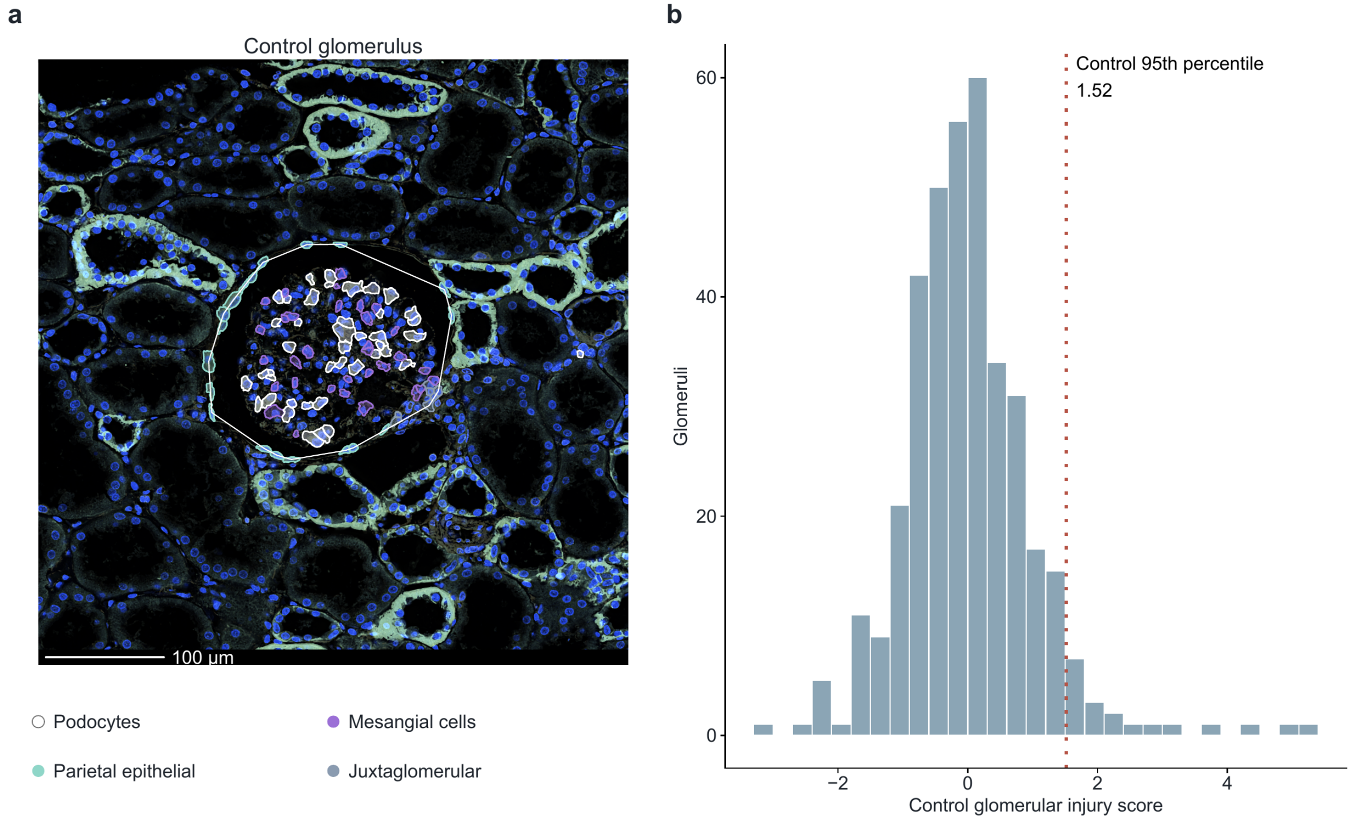


**Supplemental Figure 1.** Identification of glomeruli and the injury threshold. (a) A control field of 500 by 500 µm with the Xenium morphology image, showing podocyte, mesangial, parietal epithelial and juxtaglomerular cell boundaries and the glomerular boundary; the four cell types are identified in the color legend. Scale bar, 100 µm. (b) Injury scores of all 373 control glomeruli from 24 donors, with the 95th percentile (1.52) that defines an injured glomerulus.

#### Supplemental Figure 2

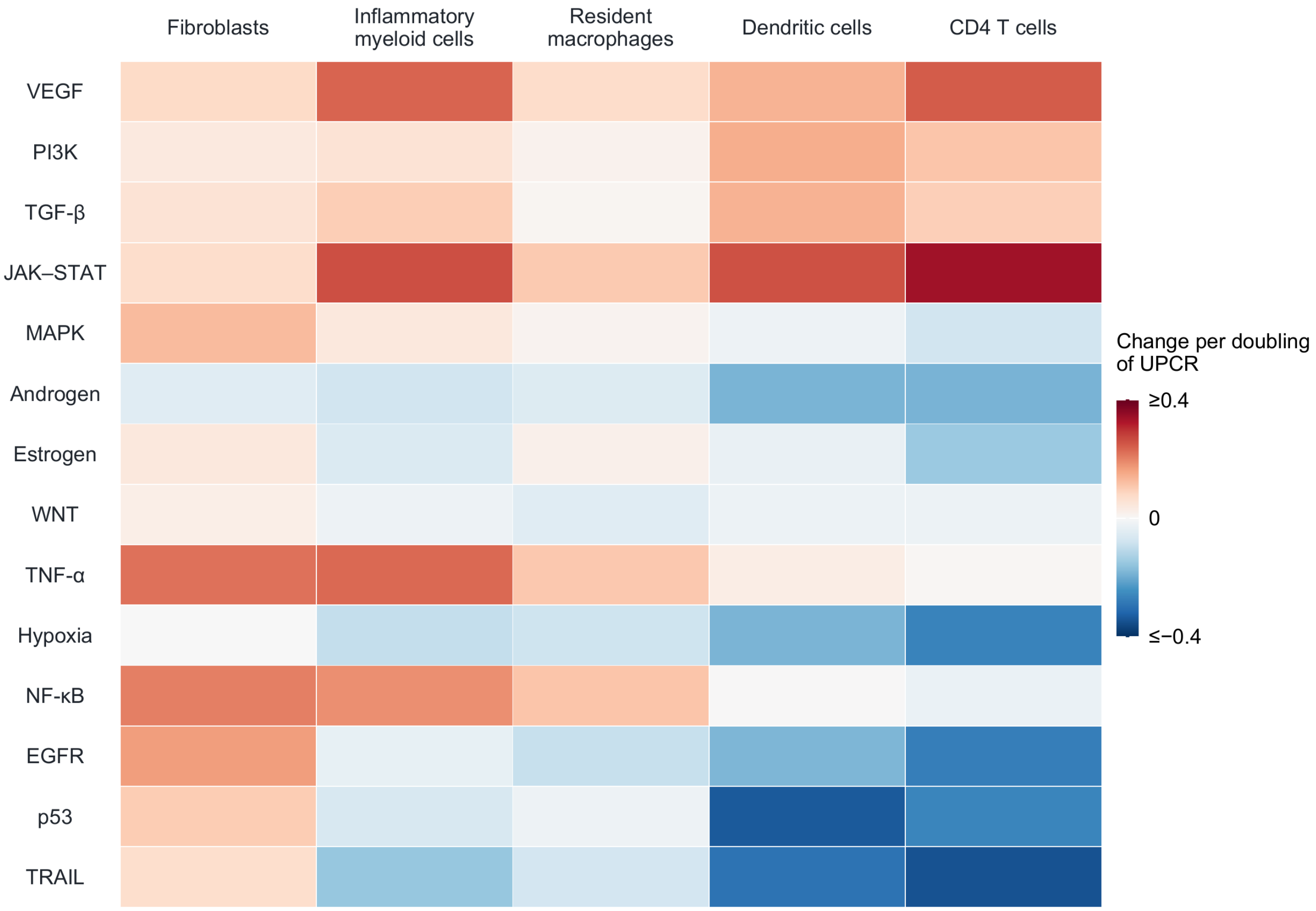


**Supplemental Figure 2.** Proteinuria and signaling in cells around proximal tubules. Change in the activity of each of 14 signaling pathways per doubling of UPCR in fibroblasts, inflammatory myeloid cells, resident macrophages, dendritic cells and CD4 T cells within 80 µm of proximal tubules, from the joint clinical model of Table 2. Analyses included 60 patients for fibroblasts, inflammatory myeloid cells and resident macrophages, 58 for dendritic cells and 59 for CD4 T cells. Confidence intervals and *P* values are given in Supplemental Data 1.
